# A multidomain intrinsic capacity score tracks longitudinal health trajectories in the UK Biobank

**DOI:** 10.64898/2026.04.10.26350621

**Authors:** Ting Zhai, Mohan Babu, Matías Fuentealba, Saleem A Al Dajani, Vadim N Gladyshev, David Furman, Michael P Snyder

## Abstract

Quantitative measures for tracking functional health have generally been lacking. Intrinsic capacity (IC) has been proposed as an appropriate measure, but its metrics have been derived in small datasets and sparse longitudinal data. Using harmonized measures of cognition, locomotion, sensory function, vitality, and psychological well-being from 501,615 UK Biobank participants and followed for a median of 15.5 years, we derived domain-specific and composite IC scores. We examined associations with incident disease, cause-specific mortality, multimorbidity, lifestyle and socioeconomic factors, and multi-omic profiles from Olink proteomics, NMR metabolomics, clinical biochemistry, and blood-cell traits. We found that composite IC declined non-linearly with age, and within-person decline was steeper than the cross-sectional age measures. Participants with greater baseline morbidity, those who subsequently developed incident disease, and those who died earlier in follow-up showed lower IC trajectories across adulthood. The IC domains were only modestly correlated with one another, supporting multidimensionality, yet higher overall IC was associated with lower risk of most diseases examined. The dominant IC domain varied by endpoint, with cognition informative for dementia, sensory function for hearing loss, psychological capacity for depression, locomotion for osteoarthritis, and vitality for cardiometabolic outcomes. IC was also associated cross-sectionally with physical activity, insomnia, smoking, medication burden, and socioeconomic disadvantage. More proteins were found predictive for vitality, and enrichment converged on immune/inflammatory and metabolic pathways. Blood-based surrogates recapitulated part of the phenotypic signal, particularly for vitality. Overall, this IC framework captures longitudinal health trajectories and broad disease vulnerability in a large middle- to older-aged cohort and supports IC as a clinically meaningful, multidomain phenotype of aging and identifies blood-based correlates that may facilitate at-scale future monitoring of aging-related function declines.

## Introduction

Population aging demands a shift from disease-centered care toward early detection of declining reserve and function. The World Health Organization (WHO) has proposed intrinsic capacity (IC) as the composite of all the physical and mental capacities that an individual can draw on; influenced by genetics and environmental conditions, IC represents functional ability and therefore what people are able to do in older age^1,2^. This framing is important because the early stages of aging are often disease-free, yet manifest as a gradual erosion across attributes such as cognition, mobility, sensory function, energy regulation, and mental well-being.

IC has now been examined across multiple aging cohorts and has shown consistent prognostic relevance for outcomes such as functional decline, disability, and mortality^3–8^. At the same time, the field remains methodologically unsettled. Existing studies have constructed IC using heterogeneous indicator sets and different scoring strategies: some deriving latent scores from statistical models, others using simpler weighted or unweighted composites, and this heterogeneity has complicated comparisons across cohorts and settings^9–12^. A recent meta-analysis reinforced both points: lower IC was associated with subsequent functional decline and mortality, but substantial between-study heterogeneity remained, underscoring the need for more comparable and better-characterized implementations^6^. The biological basis of IC is also incompletely defined. Although recent work has begun to derive blood-based molecular predictors of IC, most studies have focused on clinical operationalization and outcome prediction rather than integrating multidomain phenotyping with circulating molecular profiles^13,14^.

The UK Biobank (UKB) represents a unique resource for addressing these gaps. Previous UKB studies showed that an IC score can be constructed and that lower IC predicts adverse outcomes, but these efforts have generally relied on relatively restricted indicator sets or simplified domain representations. A large proportion of studies have been using a 4-domain score construct excluding cognition^8,15,16^, and a more recent study used ten variables for 5 domains but was restricted to a small subset due to test incompleteness^7^. However, a comprehensive study is still lacking that takes full advantage of UKB which simultaneously combines richer domain mapping, repeated assessments, broad disease ascertainment, and multi-omic characterization at biobank scale.

Here we develop a comprehensive IC framework in UK Biobank that pursues four aims. First, we derive domain-specific and composite IC scores from harmonized clinical and questionnaire data spanning the five WHO domains. Second, we characterize age-related IC trajectories using repeated measurements and determine whether those trajectories separate participants by baseline morbidity, future incident disease, and mortality. Third, we test prospective associations with a broad set of incident diseases, cancers, and cause-specific mortality, while examining whether the hypothesized primary domain is most informative for domain-concordant outcomes. Fourth, we integrate proteomic, metabolomic, biochemical, and hematological data to identify circulating correlates of IC, evaluate blood-based surrogate scores, and investigate whether baseline molecular profiles carry information about later IC decline.

## Results

### Study population and IC score framework

The overall UKB study design is shown in **Fig. 1a**. The baseline analytical sample comprised 501,615 participants (mean age 56.5 ± 8.1 years, 54.4% female). The cohort was predominantly White (94.1%), with a median Townsend deprivation index of −2.1 (interquartile range [IQR]: −3.6 to 0.6). Mean body mass index (BMI) was 27.4 ± 4.8 kg/m², and 10.6% were current smokers. Over a median follow-up of 15.5 years (IQR: 14.6–16.2), 56,495 participants (11.3%) died. The longitudinal subset included 28,927 participants with repeated assessments (**Table 1**).

**Figure 1.**
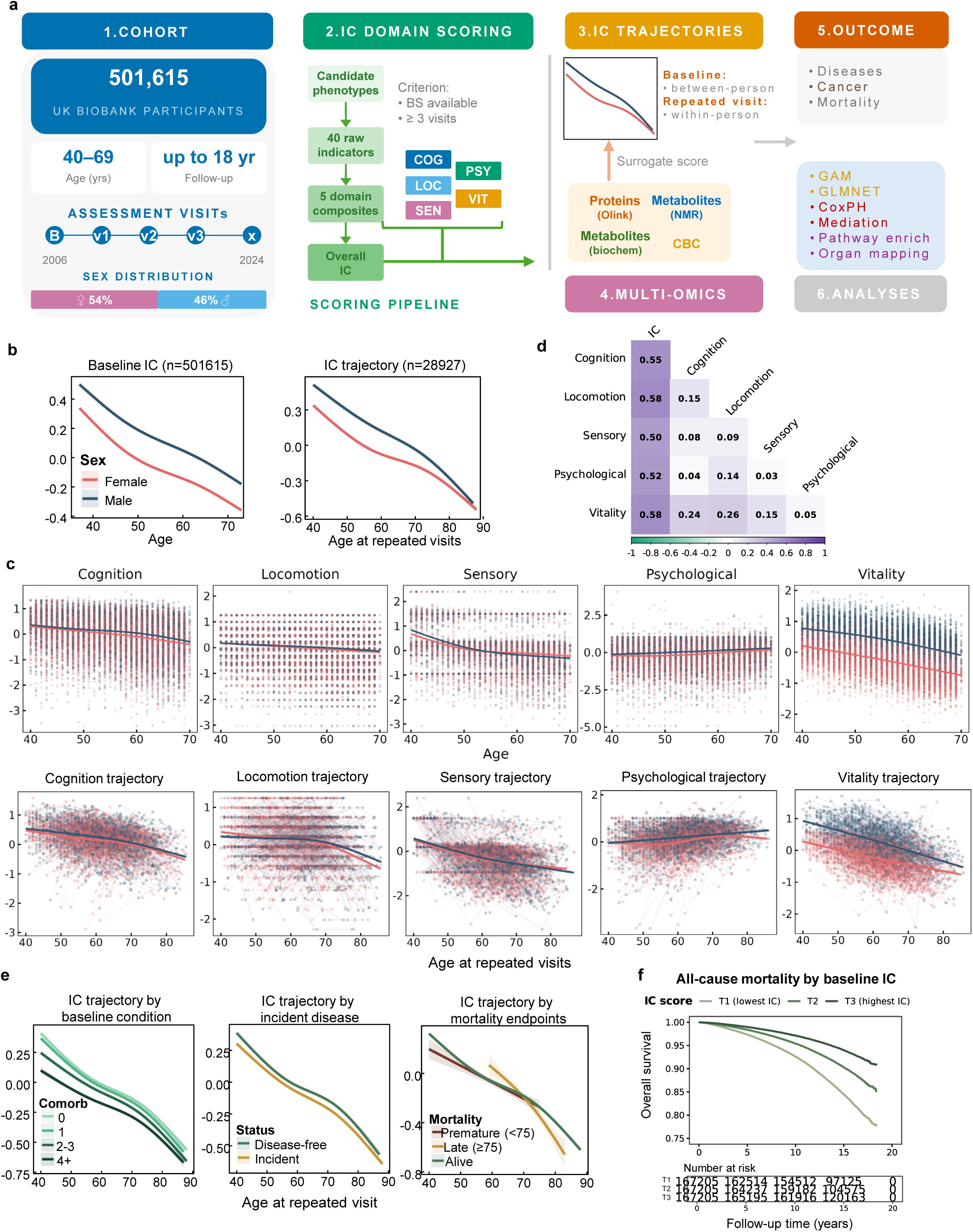
Intrinsic capacity trajectories, inter-domain structure, and mortality association. (**a**). Study design. The UK Biobank cohort was assessed at up to four visits between 2006 and 2024: baseline (B; 2006–2010), first repeat (V1; 2012–2013), first imaging (V2; 2014+), and second imaging (V3; 2019+) visits; the crossed symbol (×) denotes the administrative censoring date. IC scores were derived from 40 raw indicators aggregated into five domain composites: COG (cognition), LOC (locomotion), PSY (psychological), SEN (sensory), VIT (vitality), and an overall IC score. Baseline cross-sectional and repeated-visit longitudinal trajectories were examined alongside blood-based surrogate scores constructed from four omic platforms: Olink proteomics, NMR metabolomics, clinical biochemistry, and complete blood count (CBC). Downstream analyses comprised generalized additive models (GAM) for trajectories, elastic net regression (GLMNET) for surrogate construction, Cox proportional hazards models (CoxPH) for disease and mortality associations, causal mediation analysis, and gene-set enrichment with tissue-expression mapping. BS, baseline sample available. (**b**) Overall IC score by age at baseline (n = 501,615; left) and by age at repeated visits (n = 28,927; right), stratified by sex. Lines represent GAM-fitted smooth curves with 95% confidence intervals. (**c**) Domain-specific baseline distributions (top row) and longitudinal trajectories (bottom row) for cognition, locomotion, sensory, psychological, and vitality, by sex. (**d**) Pairwise Pearson correlation matrix among the five domain scores and the composite IC score. Inter-domain correlations are low (r = 0.03–0.26), supporting discriminant validity. (**e**) Longitudinal IC trajectories stratified by: baseline comorbidity count (0, 1, 2–3, 4+; left), incident disease status during follow-up (disease-free vs. incident; middle), and mortality outcome (premature death <75, late death ≥75, alive; right). (**f**) Kaplan–Meier curves for all-cause mortality stratified by baseline IC tertiles (T1 = lowest, T3 = highest), with number at risk.

**Table 1.** Baseline characteristics of UK Biobank participants.

| Characteristic | Overall<br>(N = 501,615) | Female<br>(N = 272,892) | Male<br>(N = 228,723) |
| --- | --- | --- | --- |
| **Sociodemographic** |  |  |  |
| **N** | 501,615 | 272,892 | 228,723 |
| **Age at recruitment (years)** | 56.5 ± 8.1 | 56.4 ± 8.0 | 56.7 ± 8.2 |
| **Female** | 272,892 (54.4%) | — | — |
| **White** | 472,173 (94.1%) | 257,122 (94.2%) | 215,051 (94.0%) |
| **Townsend deprivation index** | -2.1 (-3.6–0.6) | -2.1 (-3.6–0.5) | -2.1 (-3.6–0.6) |
| **College/university degree** | 56,798 (11.3%) | 28,436 (10.4%) | 28,362 (12.4%) |
| **Smoking status** |  |  |  |
| **Never** | 273,220 (54.5%) | 161,852 (59.3%) | 111,368 (48.7%) |
| **Previous** | 172,839 (34.5%) | 85,329 (31.3%) | 87,510 (38.3%) |
| **Current** | 52,929 (10.6%) | 24,343 (8.9%) | 28,586 (12.5%) |
| **BMI category** |  |  |  |
| **Underweight (<18.5)** | 2,624 (0.5%) | 2,077 (0.8%) | 547 (0.2%) |
| **Normal (18.5–24.9)** | 162,184 (32.3%) | 105,510 (38.7%) | 56,674 (24.8%) |
| **Overweight (25–29.9)** | 211,888 (42.2%) | 99,745 (36.6%) | 112,143 (49.0%) |
| **Obese (≥30)** | 122,137 (24.3%) | 64,246 (23.5%) | 57,891 (25.3%) |
| **Intrinsic capacity domain scores (baseline)** |  |  |  |
| **Cognition score** | -0.00 ± 0.7 | -0.05 ± 0.7 | 0.06 ± 0.7 |
| **Locomotion score** | -0.01 ± 0.7 | -0.02 ± 0.7 | 0.01 ± 0.6 |
| **Psychological score** | -0.01 ± 0.8 | -0.09 ± 0.8 | 0.09 ± 0.8 |
| **Sensory score** | 0.00 ± 0.7 | 0.02 ± 0.7 | -0.01 ± 0.8 |
| **Vitality score** | -0.00 ± 0.6 | -0.29 ± 0.5 | 0.35 ± 0.5 |
| **IC overall score** | -0.00 ± 0.4 | -0.09 ± 0.4 | 0.10 ± 0.4 |
| **Follow-up and outcomes** |  |  |  |
| **Follow-up duration (years)** | 15.5 (14.6–16.2) | 15.5 (14.7–16.2) | 15.4 (14.5–16.2) |
| **All-cause mortality** | 56,495 (11.3%) | 23,473 (8.7%) | 33,022 (14.5%) |

Values are mean ± SD for normally distributed continuous variables, median (IQR) for skewed variables, and N (%) for categorical variables. IC domain scores are standardized (z-scored) with higher values indicating greater capacity.

IC was derived from harmonized measures spanning cognition (5 cognitive tests spanning executive function, memory, and processing speed subdomains), locomotion (3 self-reported mobility items), sensory function (4 indicators from hearing and vision subdomains), vitality (7 physiological measures across muscle, respiratory, adiposity, and circulatory subdomains), and psychological capacity (3 composite scales aggregated from 21 questionnaire items reflecting distress, emotional instability, and subjective wellbeing). The full field mapping, aggregation rules, transformations, and visit availability are provided in **Supplementary Table 1**; and **Supplementary Fig. 1** shows raw or intermediate indicator distributions by age and sex. Within each domain, indicators were standardized, reverse-coded where necessary so that higher values indicate greater capacity, and averaged into unweighted domain composites as an overall IC measure. We also fitted a bifactor confirmatory factor analysis, which uses model-derived weights rather than equal averaging and is widely used in prior IC literature, and yielded similar c-index for domain-specific disease prediction (**Supplementary Table 2**).

### IC declines with age, but domains behave differently

The overall IC score showed a non-linear age gradient at baseline in both sexes. Piecewise regression on the full cross-sectional sample identified two turning points in the age–IC relationship: for females at 51.6 years (standard error [SE] 0.23) and 63.2 years (SE 0.39), and for males at 51.7 years (SE 0.37) and 61.8 years (SE 0.49), delineating three phases of initial decline, attenuated decline, and re-accelerated decline after the early 60s (**Fig. 1b**). Males had slightly higher IC scores in mid-life but showed more rapid late-life decline, such that the sex difference narrowed by age 70. In the repeated-measures subset (n = 28,927; median 3 visits over 13.7 years of follow-up), within-person IC decline averaged −0.019 standard deviation (SD) per year (total change −0.25 SD), appearing steeper than the cross-sectional gradient, consistent with cross-sectional comparisons underestimating the pace of individual-level decline owing to inter-individual heterogeneity.

Domain-specific trajectories (**Fig. 1c**) revealed substantial heterogeneity. Cognition, locomotion, sensory capacity, and vitality all declined with age, at different rates, whereas the psychological domain was comparatively stable and slightly trended upward. Examination of the individual measures contributing to the psychological domain showed similar age-related patterns for Recent Depressive Symptoms [RDS]-4, Neuroticism-12, and Wellbeing-5 (**Supplementary Fig. 1**), indicating that the domain-level pattern was not driven by a single component.

The correlation matrix among domain scores (**Fig. 1d**) confirmed the multidimensional structure of IC. The composite IC score showed moderate correlations with each domain (r = 0.50–0.58), while inter-domain correlations were generally low (r = 0.03–0.26), supporting the independent information contributed by each domain. Vitality showed the highest inter-domain correlations, particularly with locomotion (r = 0.24), consistent with partial overlap in the physiological processes captured by these domains.

Together, these results indicate that age-related decline in IC is non-linear and arises from heterogeneous trajectories across its component domains.

### IC trajectories separate participants by health course

To assess whether IC trajectories distinguished individuals on different health paths, we analyzed longitudinal IC trajectories according to three health-outcome definitions: baseline comorbidity burden, incident disease, and premature death (**Fig. 1e**). Baseline comorbidity burden showed the clearest separation: participants with no prevalent comorbidity had the highest IC trajectories across later life, whereas those with four or more conditions started markedly lower. At age 50, the predicted composite IC score was 0.16 SD (95% confidence interval [CI], 0.16 to 0.17) in those with no conditions and −0.05 SD (95% CI, −0.07 to −0.04) in those with ≥4 conditions, a difference of 0.22 SD. This gap persisted with age, remaining 0.15 SD at age 65. Differences in decline rate were comparatively modest; the highest-burden group showed slightly attenuated within-person decline relative to the no-comorbidity group (age x burden interaction β = +0.003 SD/year, P = 6.0 × 10^−18^), suggesting a possible floor effect among participants with already depleted functional reserve.

A similar but smaller level separation was observed when participants were grouped by future incident disease. At age 50, those who remained disease-free had predicted IC 0.05 SD higher than those who later developed disease (0.14 vs 0.09 SD), whereas the rates of within-person decline were similar between groups (age x incident interaction P = 0.80). Participants who died during follow-up also tended to have lower IC trajectories than survivors, although these analyses were based on much smaller samples and shorter follow-up and should therefore be interpreted more cautiously. Across these groupings, the dominant pattern was persistent level separation rather than large differences in slope, suggesting that IC captures differences in accumulated burden and remaining functional reserve before overt clinical events occur.

Consistent with these longitudinal patterns, all-cause mortality showed a clear dose-response gradient across composite IC tertiles, with persistently worse survival in the lowest tertile over 18 years of follow-up (**Fig. 1f**). Estimated survival at 5, 10, and 15 years was 97.4%, 92.6%, and 84.5% in the lowest IC tertile, compared with 98.4%, 95.4%, and 90.3% in the middle tertile, and 99.0%, 97.1%, and 94.0% in the highest tertile.

### Higher IC predicts a broad spectrum of incident disease and cause-specific mortality

Fully adjusted Cox models showed that higher overall IC was associated with lower risk of nearly every major incident disease examined (**Fig. 2a** for domain-concordant diseases**; Supplementary Table 3** for broader spectrum of health outcomes). The strongest associations were observed for psychiatric conditions (HR = 0.18 for bipolar disorder, 0.26 for psychotic disorders, 0.32 for depression per 1-SD higher overall IC), metabolic diseases (HR = 0.36 for type 2 diabetes, 0.31 for obesity), respiratory conditions (HR = 0.39 for COPD), and cardiometabolic endpoints (HR = 0.36 for heart failure, 0.53 for ischaemic heart disease). Chronic kidney disease (HR = 0.47) and chronic liver disease (HR = 0.41) also showed strong associations. Higher overall IC was associated with substantially lower multimorbidity incidence (IRR = 0.54; **Supplementary Table 4**). The prospective models also showed partial domain specificity rather than a strict one-domain–one-disease pattern. All associations were directionally concordant with the expected biology, for example, sensory capacity for hearing loss, psychological capacity for depression, and locomotion for osteoarthritis. However, several outcomes, particularly dementia and cardiometabolic diseases, were not dominated by a single expected domain. Instead, vitality emerged as the most consistently informative individual domain across a wide range of endpoints, while the overall IC score generally exceeded any single-domain association. This pattern suggests that IC captures both domain-relevant deficits and broader multisystem reserve, with vitality indexing a particularly central axis of systemic vulnerability.

**Figure 2.**
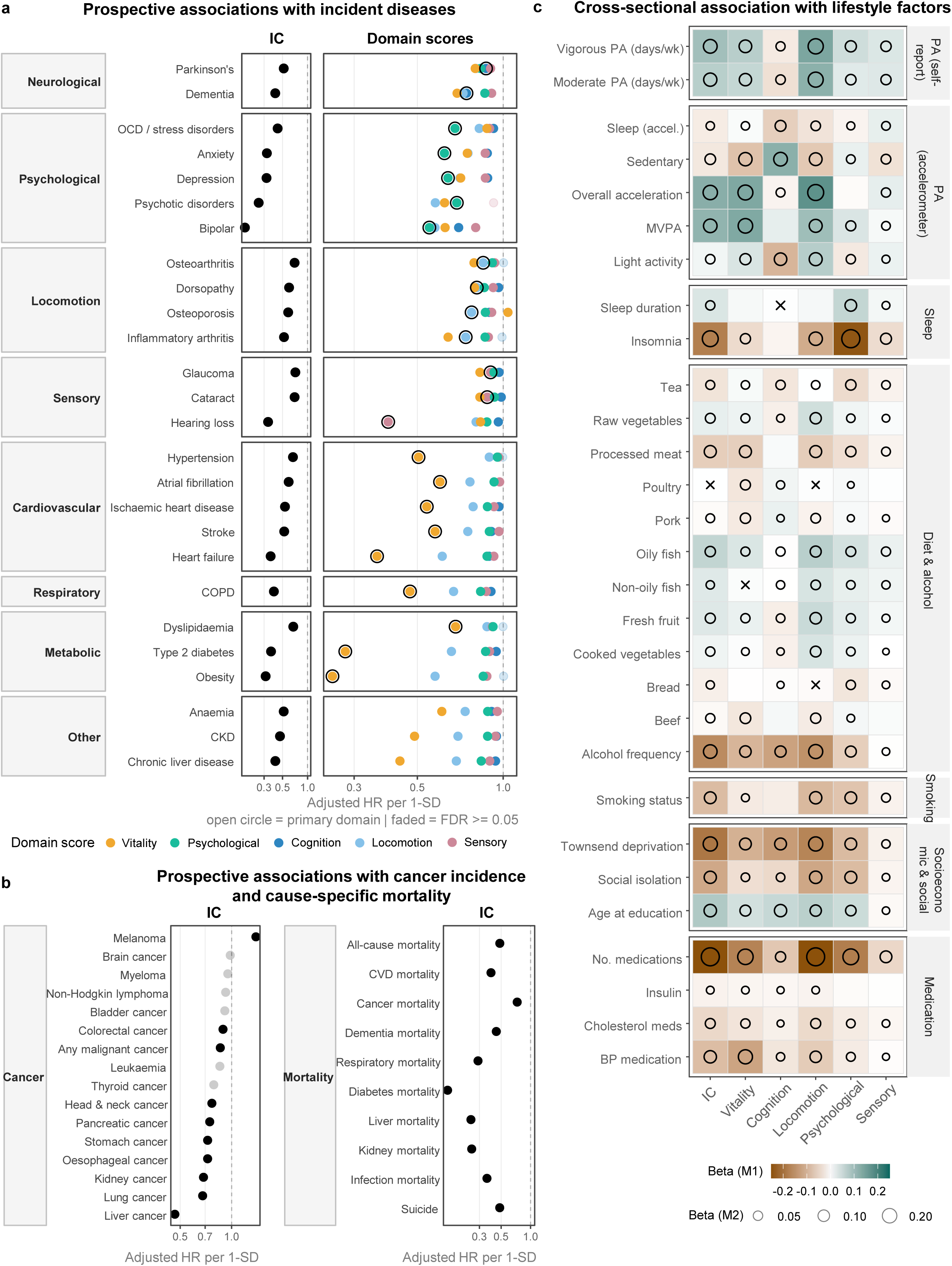
Disease associations, domain specificity, and lifestyle correlates. (**a**) Hazard ratios (per 1-SD increase) for the composite IC score (left) and domain-specific scores (right, color-coded by domain) across 33 incident disease endpoints in eight categories. Open circles indicate the hypothesized primary domain for each endpoint; faded points indicate FDR ≥ 0.05. All models were adjusted for age, sex, ethnicity, Townsend deprivation score, educational qualifications, assessment year and season, smoking status, and alcohol intake frequency. (**b**) Hazard ratios (per 1-SD increase) for the composite IC score across cancer incidence endpoints (left) and cause-specific mortality endpoints (right). The faded points indicate FDR ≥ 0.05. (**c**) Cross-sectional associations between IC domain scores and lifestyle factors. Self-reported exposures and accelerometer-derived physical activity measures were screened against overall IC and each domain score using generalized linear models. Two adjustment levels are shown: tile fill color encodes the standardized regression coefficient (β) from Model 1 (age and sex adjusted). Open circles denote associations that remained significant (FDR < 0.05) after full confounder adjustment in Model 2 (age, sex, ethnicity, Townsend deprivation score, educational qualifications, assessment year and season, smoking status, and alcohol intake frequency), with diameter encoding effect size. Crosses (x) mark associations where the direction of effect reversed between Model 1 and Model 2. Rows are grouped by exposure category: self-reported physical activity, accelerometer-derived physical activity, sleep, diet and alcohol, smoking, socioeconomic and social factors, and medication use.

Beyond incident disease, IC was also associated with cause-specific mortality (**Fig. 2b**). Per 1-SD higher overall IC, all-cause mortality HR was 0.48 (95% CI: 0.47–0.50). The strongest associations were observed for diabetes-related mortality (HR = 0.14), kidney-related mortality (HR = 0.25), liver-related mortality (HR = 0.25), respiratory mortality (HR = 0.29), and infection-related mortality (HR = 0.36). Cardiovascular mortality (HR = 0.39), dementia-related mortality (HR = 0.45), and cancer mortality (HR = 0.73) were also significantly associated, though the cancer mortality association was more modest, consistent with the heterogeneous etiology of neoplastic disease. Notably, even mortality from external causes (HR = 0.39) and suicide (HR = 0.48) showed inverse associations with IC, likely reflecting the contribution of the psychological domain and broader socioeconomic disadvantage to these endpoints.

For cancer incidence (**Fig. 2b**), the overall IC score was associated with lower risk of any malignant cancer (HR = 0.86), with the strongest site-specific associations for liver cancer (HR = 0.47), lung cancer (HR = 0.68), stomach cancer (HR = 0.72), oesophageal cancer (HR = 0.72), pancreatic cancer (HR = 0.74), head and neck cancer (HR = 0.77), kidney cancer (HR = 0.68), and uterine cancer (HR = 0.65). These cancers share strong lifestyle and metabolic risk factor profiles (smoking, adiposity, alcohol), consistent with the vitality and lifestyle components of IC. Conversely, melanoma showed a positive association (HR = 1.39), while prostate cancer showed no association after full adjustment (HR = 1.00), suggesting the modest M1 association was confounded by health-seeking behavior.

In sensitivity analyses comparing associations across different covariate adjustment strategies (**Supplementary Table 3**), associations were attenuated from M1 (age and sex) through M2 (fully adjusted) to M3 (additionally adjusting for prevalent comorbidity count), but remained significant and directionally consistent across all three models, indicating that IC retains predictive value for incident disease and mortality beyond established confounders and existing disease burden.

Sex-stratified Cox models (**Supplementary Fig. 3; Supplementary Table 5**) revealed that the inverse associations between IC and disease risk were usually present in both sexes but differed in magnitude. The largest sex differences appeared in cardiovascular endpoints (the overall IC score was more strongly protective in females for ischaemic heart disease, hypertension, and atrial fibrillation), osteoporosis (more protective in males, likely reflecting the lower baseline prevalence), and psychiatric conditions (stronger effects in males). Cancer associations were generally modest in both sexes, with the exception of lung and liver cancer. For cause-specific mortality, the protective association of IC was broadly similar in females and males across most endpoints, with the largest divergences seen for infection mortality (stronger in males) and diabetes mortality (stronger in females). Higher IC was associated with lower multimorbidity incidence in both sexes (**Supplementary Table 4**).

### Cross-sectional associations with lifestyle factors

**Fig. 2c** displays age-and-sex-adjusted (M1) and fully adjusted (M2) associations between IC scores and a comprehensive panel of lifestyle, dietary, sleep, smoking, socioeconomic, and medication variables (full results in **Supplementary Table 6**). Socioeconomic and behavioral exposures attenuated by 30–40% from M1 to M2, suggesting partial confounding among these correlated factors. Physical activity associations were stable or slightly strengthened after full adjustment. Effect directions were consistent across both adjustment strategies for all strongly associated variables.

Number of medications showed the strongest association with the overall IC score (M1 β = −0.27; M2 β = −0.23), consistent with medication burden as a proxy for multi-system disease load, followed by insomnia (M1 β = −0.18; M2 β = −0.16), which showed its largest domain-specific effect in the psychological domain (M2 β = −0.23). Townsend deprivation (a measure of socioeconomic status; M1 β = −0.19; M2 β = −0.12) and social isolation (M1 β = −0.12; M2 β = −0.07) were inversely associated across all domains, indicating the socioeconomic gradient in IC.

The locomotion domain showed the strongest positive associations with self-reported vigorous and moderate physical activity (M2 β = 0.14 for both), with concordant associations in the accelerometer-measured subsample; the vitality domain was also positively associated with physical activity, though with smaller effect sizes (M2 β = 0.07–0.08). Smoking was inversely associated with all domains, most strongly with locomotion (M1 β = −0.10; M2 β = −0.07). Dietary associations were modest in magnitude (|β| < 0.07): fruit, vegetable, and oily fish intake were positively associated with overall IC, while processed meat showed the largest negative dietary association (M2 β = −0.07). The cognition domain showed weak inverse associations with physical activity and healthy dietary variables, and a positive association with accelerometer-measured sedentary time (M2 β = 0.10), likely reflecting selection bias through occupation, as UKB cognitive assessments are speed-based touchscreen tasks that favor participants in sedentary, cognitively demanding roles^17^.

### Proteomic signal is strongest for vitality and converges on inflammation and metabolism

To identify molecular markers for IC domain we analyzed proteomic, metabolomic and clinical features. Among the four molecular platforms, proteomics yielded the richest set of IC-associated features, with vitality showing by far the largest signal (734 significant features; **Fig. 3a**; **Supplementary Table 7**). Venn diagrams (**Fig. 3b**) showed that the majority of protein-significant markers were unique to a single domain (550 proteins, 60.3%, unique to vitality), with modest cross-domain overlap. Metabolomics showed greater proportional overlap, consistent with the broader metabolic nature of metabolite features. Elastic-net reconstruction of phenotypic IC from the proteomic platform was strongest for vitality (cross-validated r^2^ = 0.72) and moderate for overall IC (r^2^ = 0.34), with weaker performance for cognition, sensory, and psychological capacity (**Supplementary Table 8**).

**Figure 3.**
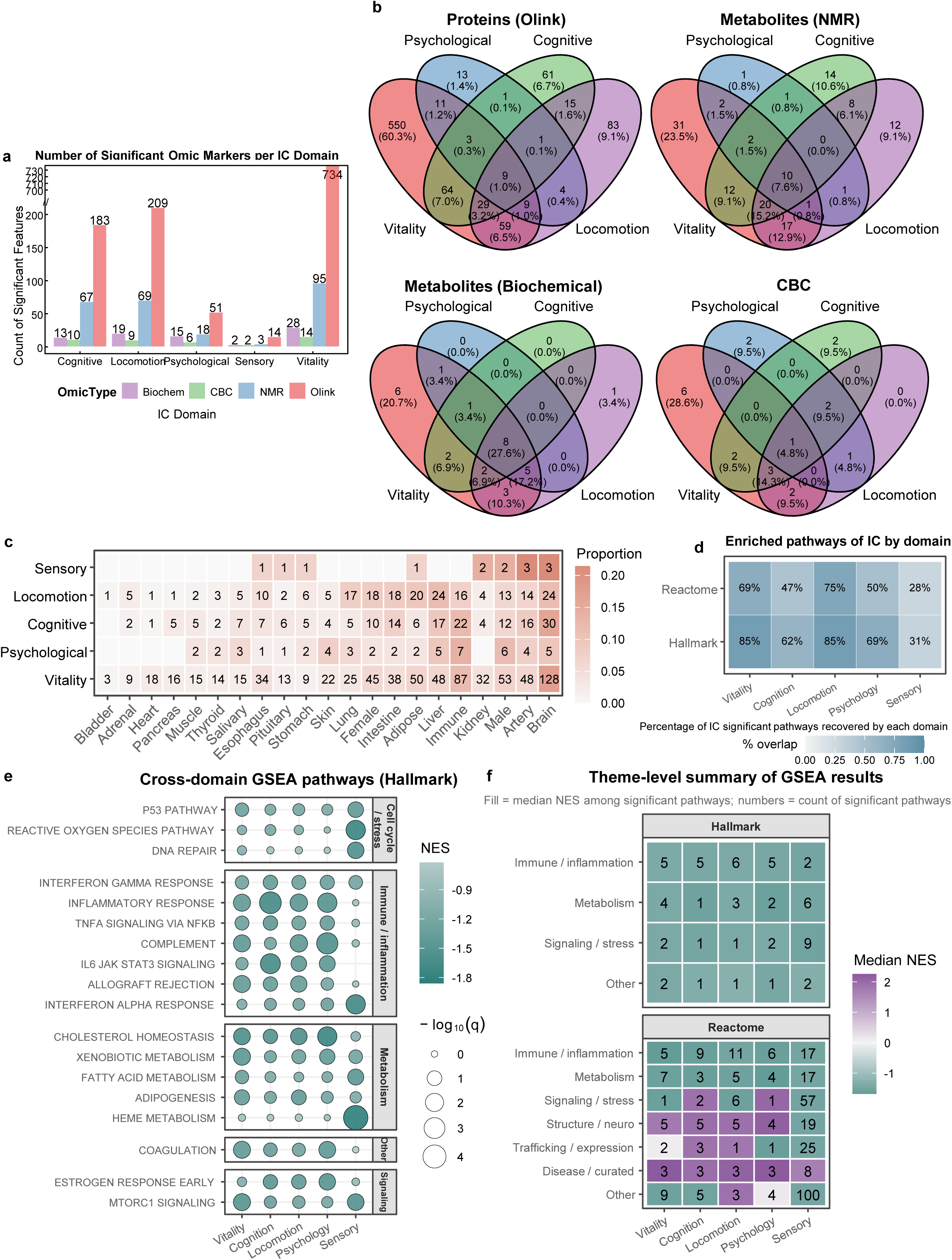
Omic surrogate markers, pathway enrichment, and tissue expression. (**a**) Stacked bar chart of significant omic marker counts per IC domain by platform. (**b**) Venn diagrams showing the overlap of significantly associated omic markers across IC domains, separately for proteins (Olink), metabolites (NMR), metabolites (biochemistry), and complete blood count (CBC) platforms. Sensory was excluded from this plot due to sparse signals. (**c**) Heatmap of the proportional tissue/organ composition of elastic net-selected proteins per IC domain. (**d**) Percentage of significant pathways of the overall IC score recovered by each individual domain, for Hallmark and Reactome gene sets. (**e**) Cross-domain Gene Set Enrichment Analysis (GSEA) bubble plot for Hallmark pathways. Dot size encodes −log10(q-value); fill color encodes normalized enrichment score (NES). (**f**) Theme-level summary of GSEA results: fill color = median NES among significant pathways; numbers = count of significant pathways per theme per domain.

Tissue-enrichment patterns (**Fig. 3c**) were directionally coherent, with cognition-associated proteins tending to be brain-enriched and vitality-associated proteins mapping more strongly to liver, adipose, lung, and kidney. Across domains, pathway analyses (**Fig. 3e,f**; **Supplementary Table 9**) repeatedly implicated immune/inflammatory signaling and metabolic programs, including interferon response, TNF–NFκB, complement, IL6–JAK–STAT3, cholesterol homeostasis, fatty-acid metabolism, and adipogenesis. Negative enrichment scores indicated that higher pathway protein expression was associated with lower domain scores. The cognitive domain showed additional enrichment for p53 and DNA repair pathways, whereas vitality was distinguished by reactive oxygen species pathway enrichment. The enriched pathway recovery analysis (**Fig. 3d**) showed that vitality recovered 85% of Hallmark and 69% of Reactome pathways significant at the composite IC level, making it the most representative single domain for the composite’s circulating biology.

### Blood-based surrogates capture part of the IC signal on future diseases

To evaluate whether blood-based surrogates capture part of the IC signal relevant to future disease, we performed two complementary analyses. Causal mediation was first performed to decompose each IC–disease association into a direct effect and an indirect effect transmitted through the matched omic surrogate, quantified as the proportion mediated. Second, we compared the incremental discrimination (cross-validated ΔC-index) of phenotype-derived IC scores, omic surrogates, and their combination over an age-and-sex baseline. Both analyses were conducted for the overall IC score and each of the five domain scores; **Fig. 4** presents overall IC results, with domain-level results in **Supplementary Tables 10,11**.

**Figure 4.**
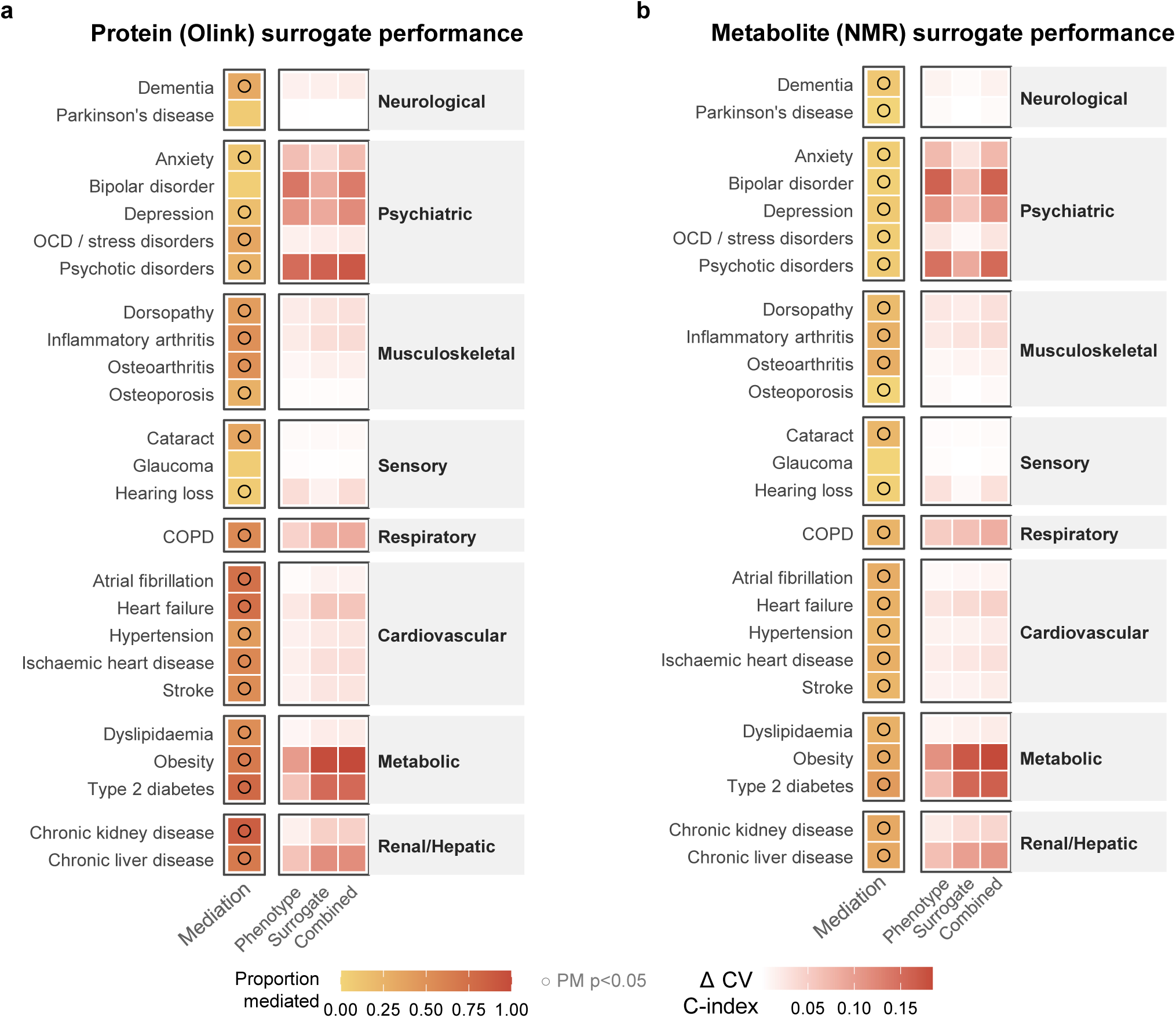
Mediation by omic surrogates and incremental discrimination. (**a**) Protein (Olink) surrogate performance for the overall IC score across incident disease endpoints. Left column: proportion of the IC–disease association mediated by the matched Olink surrogate score; open circles indicate significant mediation (p < 0.05). Right columns: cross-validated ΔC-index (incremental discrimination over age and sex) for three models: phenotype IC score alone, Olink surrogate score alone, and combined. (**b**) Corresponding panel for metabolite (NMR) surrogates.

Mediation analyses revealed that matched blood-based surrogates attenuated IC–disease associations most strongly for cardiometabolic outcomes: for example, the protein surrogate mediated over 50% of the overall IC association with obesity and type 2 diabetes, and more weakly for sensory and psychiatric endpoints (**Fig. 4a**; **Supplementary Table 10**). Metabolite surrogates showed a broadly similar pattern, though with generally lower proportions mediated and fewer endpoints reaching significance (**Fig. 4b**), consistent with the metabolomic panel capturing a narrower slice of the IC-relevant biology than the proteomic panel. At the domain level, the vitality surrogate showed particularly high mediation for metabolic and renal outcomes, while the cognition surrogate captured a substantial proportion of the dementia association (**Supplementary Table 10**). The same surrogates improved discrimination beyond age-and-sex-only models for several outcomes, especially metabolic, respiratory, and psychiatric disease, with protein surrogates generally providing larger ΔC-index gains than metabolites. Domain-level analyses revealed that the psychological domain’s phenotypic score provided substantial incremental discrimination for psychiatric outcomes (e.g., ΔC = 0.138 for bipolar disorder), while the proteomic surrogate added complementary information (**Supplementary Table 11**).

### Baseline molecular profiles contain information about subsequent IC change

In participants with at least three visits, we tested whether baseline omic surrogate scores predicted the rate of IC decline beyond the phenotypic starting point using linear mixed effect models (**Supplementary Table 12**). Overall IC decline was associated with baseline proteomic surrogate (β_interaction_ = −0.022 SD/year per SD surrogate, FDR < 0.001) and metabolite surrogate (β = −0.008, FDR < 0.001). The vitality domain showed a significant protein surrogate interaction (β = −0.027, FDR = 0.002), and the psychological (β_protein_ = −0.057, FDR < 0.001; β_metabolite_ = −0.024, FDR < 0.001) and sensory domains showed significant interactions for both platforms (β_protein_ = −0.053, FDR = 0.005; β_metabolite_ = −0.016, FDR = 0.003). The locomotion domain showed a positive protein surrogate interaction (β = 0.036, FDR = 0.006), indicating that a higher proteomic surrogate was associated with less steep locomotion decline. The cognition domain was not significantly associated with either platform. Thus, baseline omic surrogates were associated with subsequent IC slope for the composite score and for selected domains, although the direction of association differed across domains.

## Discussion

Our study supports intrinsic capacity as a scalable, multidomain phenotype of aging that is informative longitudinally, clinically relevant across a wide range of incident diseases, and linked to measurable circulating molecular correlates. Rather than functioning as a single frailty-like summary, IC captured heterogeneous patterns of reserve across domains, separated participants by subsequent health trajectory years before overt clinical events, and showed convergent biological signal in blood-based omics data. Together, these findings position IC as a useful framework for studying aging as an evolving, multisystem process rather than a late-stage accumulation of disease.

Our results demonstrate that longitudinal trajectories can serve as a summary of aging reserve. Both the baseline cross-sectional age gradient and the longitudinal within-individual analyses showed non-linear age-related patterns in IC. This non-linearity likely reflects, at least in part, the multidomain structure of IC itself: the component domains did not age in parallel, but instead showed distinct rates and shapes of change across adulthood, such that the composite score captures the combined dynamics of multiple physiological systems rather than a single homogeneous process. In the UK Biobank data, within-person IC decline was steeper than the baseline cross-sectional gradient, and trajectories for participants who later developed disease or died earlier were already shifted downward from midlife. Notably, this steeper longitudinal decline was observed despite likely positive selection into repeat assessment, which would be expected to attenuate rather than exaggerate deterioration. Together, these findings support the view that IC captures dynamic functional reserve across lifespan rather than static health status at a single time point. This aligns with the WHO framework that healthy aging should be evaluated through trajectories of function and reserve, not only through disease counts or late-stage disability^1,2^. It also supports the value of repeated IC monitoring rather than one-off screening. The observation that the composite IC trajectory emerges from heterogeneous domain-specific changes is also directionally consistent with our previous deep longitudinal profiling study, which identified turning points in multi-omic dysregulation across mid-to-late adulthood^18^. Taken together, these findings suggest that non-linear IC decline may represent the phenotypic integration of asynchronous molecular and system-level aging processes.

The disease-association patterns also strengthen the clinical interpretation of the IC domain framework. If IC were merely a relabeled marker of generalized resilience, we would expect a largely uniform rank ordering of domains across endpoints. Instead, the results support a more hierarchical structure: some outcomes aligned more closely with conceptually related domains, whereas others appeared to reflect broader multisystem decline. This pattern does not imply strict domain exclusivity, but it is consistent with the view that different IC domains capture partly distinct vulnerability pathways within an overall construct of physiological reserve. That interpretation accords with prior cohort studies in ELSA, ELSI-Brazil, and LASA showing that both composite and domain-specific IC scores predict adverse outcomes^6,10–12^; our study extends this literature through richer domain mapping, repeated measures, and broader prospective phenotyping. The cause-specific mortality analyses further emphasized the systemic nature of IC decline, with particularly strong associations for diabetes-related, renal, hepatic, and respiratory deaths.

The psychological domain warrants nuanced interpretation. In our data, its trajectory was flatter and shifted upward with age, a pattern that was also supported by the individual component measures rather than driven by a single anomalous indicator. We therefore do not regard this pattern as a simple scoring artifact. Instead, it likely reflects the fact that this domain captures dimensions of distress, neuroticism, and subjective wellbeing that do not necessarily deteriorate in parallel with physical or cognitive capacity. In population cohorts, later life may be accompanied by lower distress, better emotional regulation, response shift in self-evaluation, and selective retention of individuals with greater psychological resilience^19^. Bernal et al. reported a similar non-declining psychological trajectory using longitudinal survey data aligned to the Health and Retirement Study framework^3^, and Meng et al. identified heterogeneous trajectory classes in the Taiwan Longitudinal Study on Ageing, with some groups showing stable psychological capacity^4^. Importantly, higher psychological scores in our study still predicted lower risk of subsequent psychiatric outcomes, indicating that the domain remains clinically meaningful even though its age pattern differs from the more uniformly declining physical domains.

A distinct contribution of this study is the integration of multi-omic data into the IC framework, an aspect that has been largely absent from prior IC literature. Among the available platforms, the plasma proteome showed the richest and most consistent associations, with particularly strong signal for vitality, while pathway enrichment converged across domains on immune/inflammatory and metabolic programs. These patterns are concordant with broader aging research linking inflammaging, metabolic dysregulation, and organ-level plasma signatures to healthspan and mortality^18,20–22^, but they should be interpreted as biological correlates rather than direct mechanisms. The tissue-expression profiles add further plausibility, with domain-associated proteins mapping in expected directions to brain for cognition, musculoskeletal and connective tissues for locomotion, and liver/adipose-related signatures for vitality.

The surrogate analyses also suggest potential translational value. Mediation analyses showed that proteomic surrogates accounted for a meaningful proportion of the IC-disease association, particularly for metabolic and cardiovascular outcomes, and combined phenotype-plus-surrogate models generally provided the best discrimination. Baseline proteomic and metabolomic surrogates were also associated with subsequent IC slope for selected domains after accounting for baseline phenotypic IC, suggesting that circulating profiles may identify aspects of vulnerability not fully captured by clinical assessment alone. However, these conditional change analyses remain exploratory, and their mixed directionality across domains indicates that the biological determinants of decline may not be uniform. In particular, such estimates may also be influenced by regression to the mean and scale constraints. Accordingly, these results should not be interpreted causally, and the clinical utility of omic surrogates will require external validation before they can be considered monitoring tools.

Our study has both strengths and limitations. Strengths include the sample size (>500,000), long follow-up (median 15.5 years), repeated clinical measures (∼29,000 with ≥3 visits), comprehensive record linkage for disease ascertainment, and access to multiple molecular platforms. We also note several limitations. The UK Biobank is healthier and less diverse than the source population (94% White, higher education, lower deprivation), and the repeated-measures subset is further selected, therefore conditioning on the repeated subset might introduce selection bias, though we found similar non-linear trajectory with baseline samples. Some domains rely on self-report or proxy indicators, especially locomotion and vision, which may reduce sensitivity to early decline. The disease models remain susceptible to residual confounding, though the reported associations have been adjusted for covariates including demographic, lifestyle, and socioeconomic factors. Analyses are observational and external validation would be needed.

### Future implications and translational relevance

These findings have implications for both measuring aging and future implementation. First, IC appears to be measurable at scale in population cohorts using routinely collected clinical and questionnaire data, supporting its use as an intermediate functional phenotype between molecular aging and overt disease. Second, the domain-specific and composite associations suggest that IC could be used both as a global monitoring tool and as a stratification framework to identify which functional systems are deteriorating most rapidly in an individual. Third, the omic surrogate analyses raise the possibility that a reduced biomarker panel could complement phenotypic assessment in settings where repeated functional testing is impractical. Finally, because IC trajectories diverged years before incident disease and mortality outcomes, future work should test whether IC can serve as an enrichment marker, intermediate endpoint, or treatment-monitoring tool in preventive trials targeting midlife and early older age. External validation in more diverse cohorts, repeated-omics designs, and intervention studies will be important next steps toward clinical translation.

## Conclusion

Our study supports IC as a useful phenotype for characterizing aging trajectories in midlife and later life. The multidomain structure, within-person trajectory separation by subsequent health course, outcome-concordant domain specificity, and convergent circulating correlates collectively support IC as a clinically meaningful construct. These findings provide a foundation for scalable monitoring of healthy aging and support IC as a measurable target for risk stratification and prevention-oriented aging research.

## Methods

### Study population

The UK Biobank is a prospective population-based cohort study that recruited 502,461 participants aged 40–69 years between 2006 and 2010 across 22 assessment centers in England, Scotland, and Wales. Participants underwent physical measurements, provided biological samples, and completed touchscreen questionnaires and cognitive tests at baseline. A subset of approximately 29,000 participants attended a repeat assessment visit (2012–2013), and further imaging assessment visits were initiated from 2014 and 2019. The study was approved by the North West Multi-Centre Research Ethics Committee, and all participants provided written informed consent. We excluded participants who withdrew consent and those with missing data on all IC domain indicators (n = 321). The final analytical sample comprised 501,615 participants for baseline IC analyses and 28,927 for longitudinal trajectory analyses.

The data were analyzed under application number 41751.

### IC domain construction

We constructed IC scores across the five WHO-defined domains using the most complete clinical measures spanning the four UKB assessment visits (2006–2010, 2012–2013, 2014+, 2019+). We distinguish the constructs in three levels: (i) raw UKB questionnaire/test fields (44 item-level fields), (ii) aggregated analytic indicators formed by combining related items into interpretable measures (21 analytic indicators), and (iii) domain-specific scores formed by z-scoring and averaging of aggregated analytic indicators within each IC domain. Full details are provided in **Supplementary Table 1**.

#### Cognitive domain

Five tests administered at all in-clinic visits were selected for longitudinal comparability^17^: reaction time (log-transformed, reverse-coded), pairs matching (reverse-coded as−ln(errors + 1)), fluid intelligence (z-scored), prospective memory (z-scored), and numeric memory (z-scored). Additional tests available only at later visits or online were excluded from the primary composite; sensitivity analyses exploring all eight indicators are reported in the **Supplementary Fig 4**.

#### Locomotion domain

Locomotor capacity was assessed using three self-reported mobility indicators: usual walking pace (ordinal 1–3; slow/steady/brisk), frequency of stair climbing in the past four weeks (ordinal 0–5), and number of falls in the past year (count, reverse-coded). These indicators were selected to reflect perceived functional mobility rather than volitional physical activity behavior that was available in UKB (e.g., metabolic equivalent task scores or exercise duration), consistent with the WHO framing of IC as underlying capacity rather than lifestyle.

#### Sensory domain

Hearing was assessed using self-reported hearing difficulty and speech reception threshold in noise; and vision was evaluated using glasses/contact lens use and reasons for glasses coded as ordinal severity for refractive error. LogMAR visual acuity was excluded because it was measured in only a small subset at the first two visits and did not improve prediction of incident eye diseases.

#### Vitality domain

Seven physiological measures were integrated across four subdomains: muscle strength (mean of right- and left-hand grip strength), respiratory function (best FEV_1_, best FVC), metabolic/adiposity (waist circumference, body fat percentage; both reverse-coded), and circulatory function (systolic blood pressure, pulse pressure; both reverse-coded). All indicators were winsorised at the 0.5th/99.5th percentile prior to z-scoring.

#### Psychological domain

Three composite scales were aggregated from 21 questionnaire items following previous methodology paper^23^: RDS-4 (four items; summed and reverse-coded), neuroticism-12 (12 items; summed and reverse-coded), and a five-item wellbeing composite (happiness and satisfaction with health, family, friendships, and finances; averaged if ≥3 non-missing). Compared to the probable depression status battery which was also available across visits, these scales were chosen to reflect psychological state and traits rather than static psychiatric history.

#### Scoring

Within each domain, analytic indicators were z-scored and reverse-coded so higher values represent greater capacity. The primary domain score was computed as the unweighted mean of the standardized indicators within each domain, a formative composite approach that treats IC as an aggregate of its constituent domains rather than a reflection of a single latent trait, consistent with the predominant scoring strategy in the IC literature^12,13^. As a sensitivity analysis, we fitted a bifactor confirmatory factor analysis (CFA), a latent variable model that estimates a general IC factor shared across all domains alongside five domain-specific factors, and compared its performance with the primary composite for downstream disease association. The two approaches yielded similar results (**Supplementary Table 2**), and the unweighted composite was retained as the primary metric, which avoids assumptions about indicator weighting and for generalizability to other cohorts.

### Covariates

All covariates were assessed at baseline unless otherwise specified. Models were adjusted for age at assessment, sex, ethnic background, assessment timing (year and season), socioeconomic status (Townsend deprivation index), educational attainment, smoking status, and alcohol intake frequency. Ethnic background was categorized into White British, Other White, and Other. Educational attainment was derived from self-reported qualifications and coded as an ordered variable reflecting highest qualification achieved. Smoking status was classified as never, former, or current smoker, and alcohol intake frequency was modeled as an ordered categorical variable. Assessment season was defined using calendar months. Full variable definitions, coding schemes, and UK Biobank field mappings are provided in **Supplementary Table 13**.

### Omic data

#### Proteomics

Plasma proteomics were measured using the Olink Explore 3072 proximity extension assay. To ensure generalizability to the full UKB baseline population, we restricted analyses to the randomly selected UKB-PPP subset, excluding the consortium-selected enrichment subset (UKB-PPP index, Field 30903) which was enriched for common risk factors and could introduce collider bias into IC–protein associations^24^. Normalized Protein eXpression (NPX) values were provided by UKB on the log2 scale after Olink’s standard intensity normalization and inter-plate bridging. We applied additional participant-level and protein-level quality control, excluding individuals with >50% proteins missing and proteins with >10% missingness^21,24^. After QC, 2,916 proteins were retained across 39,887 participants.

#### Metabolomics

Plasma NMR metabolomics were quantified using the Nightingale Health platform (Category 220). Preprocessing was performed using the ukbnmr R package^25^, which applies log transformation, removes technical variation through sample-level normalization and batch-effect correction, and derives secondary biomarkers. The resulting dataset comprised 249 directly quantified metabolites and 78 derived ratios (327 features total) available in 488,088 participants at baseline.

#### Biochemistry and CBC

Clinical biochemistry (Category 17518) and complete blood count (CBC; Category 100081) were obtained from baseline blood samples. Two biochemistry assays with high missingness (oestradiol, rheumatoid factor) were excluded, and two derived markers were computed (non-albumin protein [total protein minus albumin] and AST/ALT ratio)^26^. For CBC, extreme outlier values were set to missing (white blood cell count >200 ×10⁹/L, hemoglobin >20 g/dL, hematocrit >60%, platelet count >1000 ×10⁹/L), and participants whose differential white cell percentages summed outside 80–120% were excluded. All analytes were winsorised at the 0.1st and 99.9th percentiles. CBC counts were transformed using the inverse hyperbolic sine function scaled by the within-variable median, and percentage indices via the logit transform; biochemistry analytes with highly skewed distributions (CRP, GGT, triglycerides, lipoprotein(a)) were log1p-transformed, while liver enzymes and hormones (ALT, AST, ALP, bilirubin, urate, urea, IGF-1, testosterone, SHBG) were log-transformed. All features were subsequently z-scored. Features with >20% missingness (CBC) or >30% missingness (biochemistry) were excluded.

### Health outcomes

Disease endpoints were ascertained from the algorithmically defined first-occurrence fields (Category 1712), which integrate hospital inpatient records (HES), primary care data, self-reported diagnoses from assessment center interviews, and death registrations. Each endpoint was mapped to three-character ICD-10 codes as detailed in **Supplementary Table 14**. The full analysis comprised 34 incident disease endpoints across nine clinical categories (neurological/neurodegenerative, psychiatric, musculoskeletal, sensory, respiratory, cardiovascular, metabolic, renal, and hepatic), 20 site-specific incident cancer endpoints, and 13 cause-specific mortality endpoints. Cancer diagnoses were obtained from linked national cancer registrations (Category 100092) and, where a cancer was recorded only at death, from death registrations (Category 100093). The diagnosis date was defined as the earliest cancer registration date or the date of death if no registration preceded it. Cause-specific mortality was ascertained from the primary cause of death, classified using ICD-10 chapter-level groupings. Administrative censoring dates were region-specific for each endpoint, determined based on the UKB website.

Prevalent disease was defined as a first recorded occurrence on or before the date of baseline assessment; affected participants were excluded from the corresponding endpoint-specific analysis to ensure incident ascertainment. Comorbidity burden at baseline was quantified as the count of distinct three-character ICD-10 codes with a first recorded date on or before baseline; incident comorbidity burden was the corresponding count accruing after baseline and on or before the censoring date. Person-time was calculated from the date of baseline assessment to the earliest of: first occurrence of the endpoint of interest, death, loss to follow-up, or the applicable administrative censoring date.

### Statistical analyses

#### Longitudinal trajectories

Among participants with repeated IC measurements, age-related trajectories were estimated using generalized additive models (GAMs) with penalized cubic regression splines, with age at assessment as the smooth term and random intercepts for participant. Trajectories were stratified by sex, baseline comorbidity count (0, 1, 2–3, 4+), incident disease status (disease-free vs. incident during follow-up), and mortality outcome (alive, late death [≥75], premature death [<75]).

#### Cross-sectional lifestyle associations

We evaluated cross-sectional associations between IC scores (composite and five domain-specific scores) and a broad set of lifestyle, dietary, sleep, smoking, socioeconomic, and medication-related variables using generalized linear models. Both IC scores and continuous exposures were standardized prior to analysis, while categorical variables were modeled as ordinal predictors where appropriate. For each exposure–score pair, two models were fitted: Model 1 adjusted for age at assessment and sex, and Model 2 further adjusted for ethnic background, assessment year, assessment season, Townsend deprivation index, highest educational attainment, smoking status, and alcohol intake frequency. Accelerometer-derived physical activity measures, including overall acceleration, moderate-to-vigorous physical activity, light activity, sedentary time, and sleep duration, were available in a subsample with valid wear at instance 2. For these analyses, IC scores measured at the corresponding visit were used to align measurement timing. Covariate adjustment followed the same strategy in Model 2, with assessment year and season derived from the accelerometer wear date to reflect the timing of measurement, while other covariates were retained from baseline. Missing data were handled by complete-case analysis.

#### Disease associations

Associations between IC scores and incident outcomes were evaluated using Cox proportional hazards models, estimating HRs per 1-SD increment in each IC score. The Efron method was used to handle tied event times. For each endpoint, participants with prevalent disease at baseline were excluded to ensure prospective ascertainment.

Three models were fitted for each exposure–endpoint pair. Model 1 adjusted for age and sex. Model 2 additionally adjusted for predefined demographic and lifestyle covariates. Model 3 further included baseline comorbidity burden (count of prevalent conditions) to account for pre-existing multimorbidity. Endpoints with fewer than 50 participants or 20 events were excluded from analysis. Domain specificity was evaluated by comparing the magnitude of associations across IC domains for each endpoint. Sex-specific effects were examined using stratified models. As a sensitivity analysis, a 2-year landmark analysis was conducted for mortality outcomes, requiring participants to survive at least 2 years after baseline and left-truncating follow-up accordingly to reduce potential reverse causation.

Comorbidity burden at baseline and during follow-up was modelled as a count outcome using Poisson regression with log follow-up time as an offset. Overdispersion was assessed using the Pearson statistic, and negative binomial models were used when dispersion exceeded 1.2.

#### Omic surrogate construction

For each omic panel (proteomic [Olink], metabolomic [NMR], complete blood count [CBC], and plasma/serum biochemistry data), features were entered into elastic net regression adjusted for age and sex (unpenalized) within a nested cross-validation (CV) framework: 5-fold outer CV repeated 3 times, with 5-fold inner CV for tuning the elastic net mixing parameter (α grid: 0–0.50 for proteins, 0–0.75 for metabolites). The more regularized λ (1-SE rule) was selected throughout. Feature stability was assessed via 100 bootstrap resamples (80% subsamples), retaining features selected in ≥50% of resamples (≥60% for metabolites) with ≥80% sign consistency, followed by correlation pruning at |r| > 0.85 for proteins and > 0.90 for metabolites. Final surrogate scores were derived from a reduced elastic net refitted on the stable feature set using the full sample.

#### Mediation analysis

The proportion of the IC–disease association mediated by the matched omic surrogate was estimated using regression-based causal mediation framework^27^, with a Cox proportional hazards outcome model and a linear mediator model, adjusting for age and sex. Inference was obtained via 100 bootstrap resamples, and the proportion mediated was derived from the ratio of the pure natural indirect effect to the total effect. Endpoints with fewer than 100 participants or 20 events were excluded.

#### Risk prediction discrimination

Incremental predictive value was evaluated by 5-fold cross-validated ΔC-index relative to an age-and-sex base model. Three models were compared: (1) phenotype IC + age + sex, (2) omic surrogate + age + sex, and (3) combined.

#### Omic–trajectory interaction models

Linear mixed models were fitted among participants with ≥3 visits, separately for each IC domain and omic platform. Uncorrelated random intercepts and slopes were specified, and models were fitted with REML using the BOBYQA optimizer. The interaction term tests whether the baseline omic surrogate predicts the rate of IC change beyond the phenotypic starting point. FDR correction was applied across all models.

#### Pathway and tissue enrichment

To characterize the biological processes underlying IC–protein associations, gene set enrichment analysis (GSEA) was performed on the full proteome-wide t-statistics from linear regression models adjusted for age and sex, ranked by effect size for each IC domain, using the multilevel algorithm (gene set size 15–500) with Hallmark and Reactome gene sets from MSigDB^28^. Pathways with negative normalized enrichment scores (NES < 0) indicate enrichment among proteins associated with poorer functional status. Separately, tissue expression profiles of the elastic net–selected protein features were summarized by annotated organ category from a previous study^21^, in which genes expressed at least four-fold higher in a single organ than any other organ based on bulk RNA sequencing data from the Genotype-Tissue Expression (GTEx) Atlas were mapped to Olink-measured plasma proteins, and visualized as a proportional heatmap, with formal over-representation tests (ORA) evaluated for each domain.

#### Software

All analyses were performed in R (version 4.4.0) on the UK Biobank Research Analysis Platform. Key packages: lavaan (CFA), mgcv (GAMs), survival (Cox models), clusterProfiler (GSEA), glmnet (elastic net), CMAverse (mediation), ggplot2 (visualization).

## Data Availability

The UK Biobank data are available under controlled access. This project is approved under application number 41751. Researchers requiring access should apply (https://www.ukbiobank.ac.uk/enable-your-research/apply-for-access) and pay a fee depending on the tier of data requested.

## Code Availability

All custom code used for the analyses is available on GitHub: https://github.com/zhaiting/ukb_ic.

## Acknowledgements

We acknowledge financial support from Anu and BVJagadeesh Family Foundation.

## Author Contribution Statement

T.Z. and M.P.S. conceptualized and designed the study. M.B. managed the project. T.Z. performed the statistical analyses, visualized the data, and drafted the manuscript. M.P.S. supervised the work and revised the manuscript. M.F., S.A.A., V.N.G. and D.F. contributed to the conceptual interpretation of the results, provided scientific input and discussion. All authors reviewed and approved the final version.

## References

1. Beard, J.R., et al. The World report on ageing and health: a policy framework for healthy ageing. Lancet 387, 2145–2154 (2016).

2. World Health, O. World report on ageing and health, (World Health Organization, Geneva, 2015).

3. Bernal, M.C., Batista, E., Martinez-Balleste, A. & Solanas, A. A functional approach to model intrinsic capacity in ageing trajectories. Sci Rep 15, 7878 (2025).

4. Meng, L.C., et al. Multi-Trajectories of Intrinsic Capacity Decline and Their Impact on Age-Related Outcomes: A 20-Year National Longitudinal Cohort Study. Aging Dis 15, 2697–2709 (2023).

5. Perianayagam, A., et al. An assessment of intrinsic capacity among older Indian adults from the Longitudinal Ageing Study in India. Nat Aging 5, 2482–2493 (2025).

6. Sanchez-Sanchez, J.L., et al. Association of intrinsic capacity with functional decline and mortality in older adults: a systematic review and meta-analysis of longitudinal studies. Lancet Healthy Longev 5, e480–e492 (2024).

7. Beyene, M.B., et al. Development and validation of an intrinsic capacity score in the UK Biobank study. Maturitas 185, 107976 (2024).

8. Ramirez-Velez, R., Borda, M.G., Saez de Asteasu, M.L. & Izquierdo, M. Intrinsic capacity as a predictor of depression onset in middle-aged and older adults: Insights from the UK Biobank. J Affect Disord 388, 119590 (2025).

9. Beard, J.R., Jotheeswaran, A.T., Cesari, M. & Araujo de Carvalho, I. The structure and predictive value of intrinsic capacity in a longitudinal study of ageing. BMJ Open 9, e026119 (2019).

10. Aliberti, M.J.R., et al. Validating intrinsic capacity to measure healthy aging in an upper middle-income country: Findings from the ELSI-Brazil. Lancet Reg Health Am 12, 100284 (2022).

11. Campbell, C.L., Cadar, D., McMunn, A. & Zaninotto, P. Operationalization of Intrinsic Capacity in Older People and Its Association With Subsequent Disability, Hospital Admission and Mortality: Results From The English Longitudinal Study of Ageing. J Gerontol A Biol Sci Med Sci 78, 698–703 (2023).

12. Koivunen, K., et al. Development and validation of an intrinsic capacity composite score in the Longitudinal Aging Study Amsterdam: a formative approach. Aging Clin Exp Res 35, 815–825 (2023).

13. Beyene, M.B., Visvanathan, R. & Amare, A.T. Intrinsic Capacity and Its Biological Basis: A Scoping Review. J Frailty Aging 13, 193–202 (2024).

14. Fuentealba, M., et al. A blood-based epigenetic clock for intrinsic capacity predicts mortality and is associated with clinical, immunological and lifestyle factors. Nat Aging 5, 1207–1216 (2025).

15. Sun, M., et al. Intrinsic Capacity, Polygenic Risk Score, APOE Genotype, and Risk of Dementia: A Prospective Cohort Study Based on the UK Biobank. Neurology 102, e209452 (2024).

16. Hu, W., et al. Intrinsic Capacity Defined Using 4 Domains, Genetic Risk, and Incident Parkinson Disease: A Prospective Cohort Study. Neurology 105, e214144 (2025).

17. Fawns-Ritchie, C. & Deary, I.J. Reliability and validity of the UK Biobank cognitive tests. PLoS One 15, e0231627 (2020).

18. Shen, X., et al. Nonlinear dynamics of multi-omics profiles during human aging. Nat Aging 4, 1619–1634 (2024).

19. Stone, A.A., Schwartz, J.E., Broderick, J.E. & Deaton, A. A snapshot of the age distribution of psychological well-being in the United States. Proc Natl Acad Sci U S A 107, 9985–9990 (2010).

20. Goeminne, L.J.E., et al. Plasma protein-based organ-specific aging and mortality models unveil diseases as accelerated aging of organismal systems. Cell Metab 37, 205–222 e206 (2025).

21. Oh, H.S., et al. Plasma proteomics links brain and immune system aging with healthspan and longevity. Nat Med 31, 2703–2711 (2025).

22. Ahadi, S., et al. Personal aging markers and ageotypes revealed by deep longitudinal profiling. Nat Med 26, 83–90 (2020).

23. Dutt, R.K., et al. Mental health in the UK Biobank: A roadmap to self-report measures and neuroimaging correlates. Hum Brain Mapp 43, 816–832 (2022).

24. Sun, B.B., et al. Plasma proteomic associations with genetics and health in the UK Biobank. Nature 622, 329–338 (2023).

25. Ritchie, S.C., et al. Quality control and removal of technical variation of NMR metabolic biomarker data in ∼120,000 UK Biobank participants. Sci Data 10, 64 (2023).

26. Carrasco-Zanini, J., et al. Proteomic signatures improve risk prediction for common and rare diseases. Nat Med 30, 2489–2498 (2024).

27. Zhai, T., et al. Telomere Length Dynamics As a Biomarker of Individual Radiation Sensitivity and Pneumonitis in Lung Cancer Patients Receiving Thoracic Radiotherapy. Int J Radiat Oncol Biol Phys (2026).

28. Subramanian, A., et al. Gene set enrichment analysis: a knowledge-based approach for interpreting genome-wide expression profiles. Proc Natl Acad Sci U S A 102, 15545–15550 (2005).

