## Supplementary Fig for "A multidomain intrinsic capacity score tracks longitudinal health trajectories in the UK Biobank"

### Table of contents

**Supplementary Figure 1.** Raw indicator distributions by age and sex.

**Supplementary Figure 2.** Reactome cross-domain enrichment pathways.

**Supplementary Figure 3.** Sex-stratified hazard ratios for the composite IC score across disease, cancer, and mortality endpoints.

**Supplementary Figure 4.** Cross-sectional and longitudinal trajectories of individual cognitive test measures by assessment source.

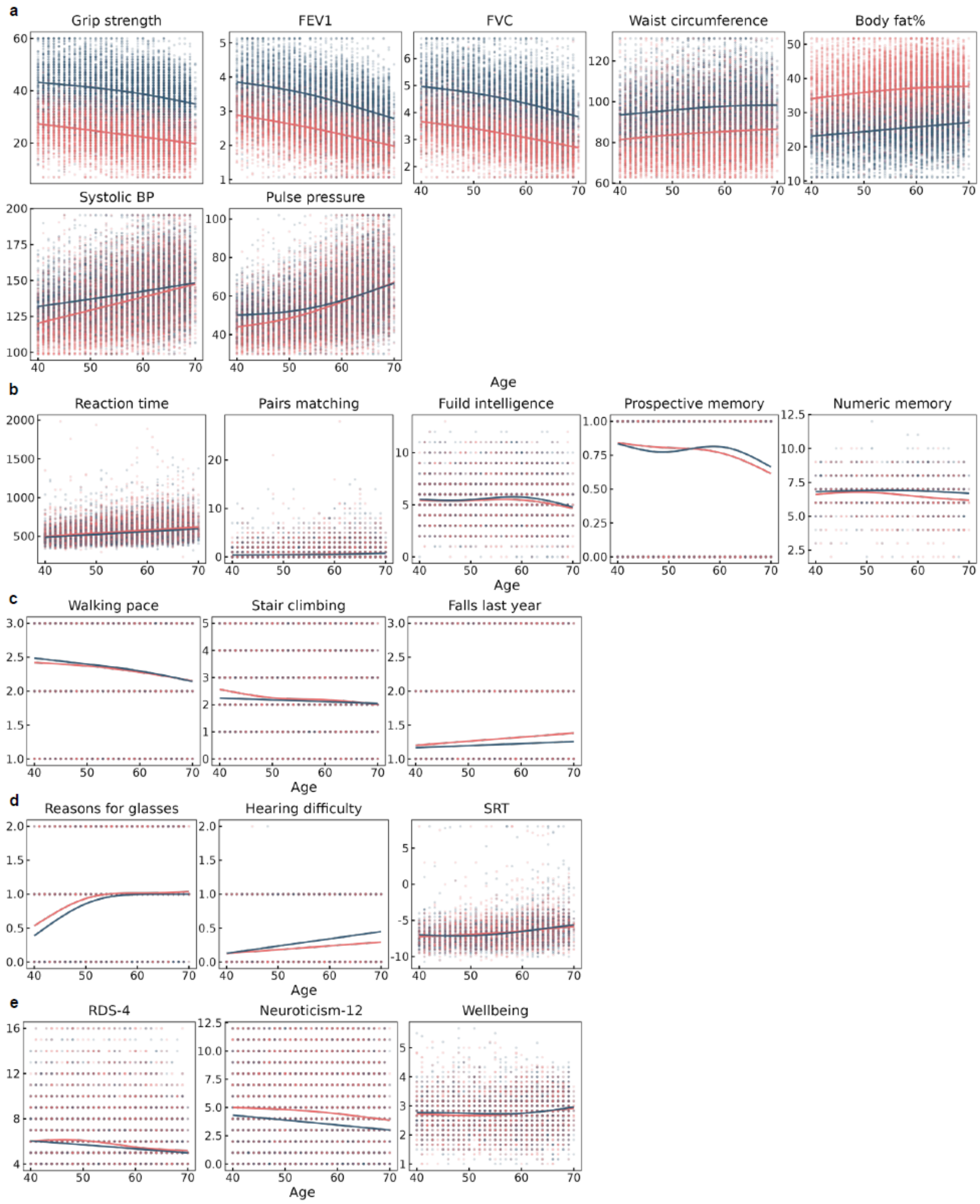


**Supplementary Figure 1. Raw indicator distributions by age and sex.** Age-stratified scatter plots with sex-stratified GAM smooth curves for all 20 raw/processed IC indicators before standardization. Indicators are grouped by domain: **a**) vitality (grip strength, FEV1, FVC, waist circumference, body fat%, systolic BP, pulse pressure), **b**) cognition (reaction time, pairs matching, fluid intelligence, prospective memory, numeric memory), **c**) locomotion (walking pace, stair climbing, falls last year), **d**) sensory (reasons for glasses, hearing difficulty, SRT), and **e**) psychological (RDS-4, neuroticism-12, wellbeing).

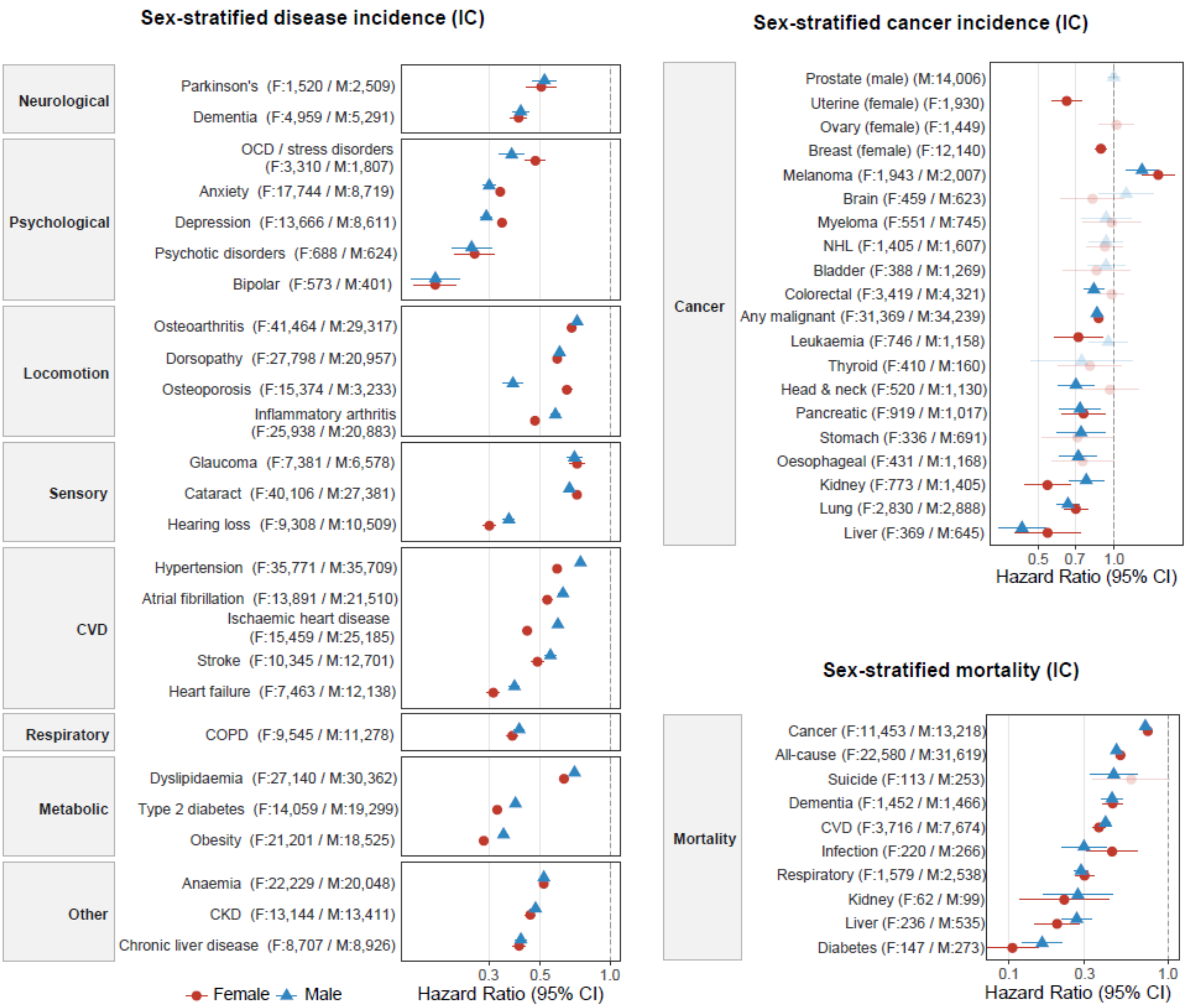


**Supplementary Figure 2. Reactome cross-domain enrichment pathways.** Extended Reactome pathway bubble plot for all five IC domains, showing disease/curated, ECM/structure, immune/inflammation, metabolism, neuro/synaptic, and signaling pathway categories.

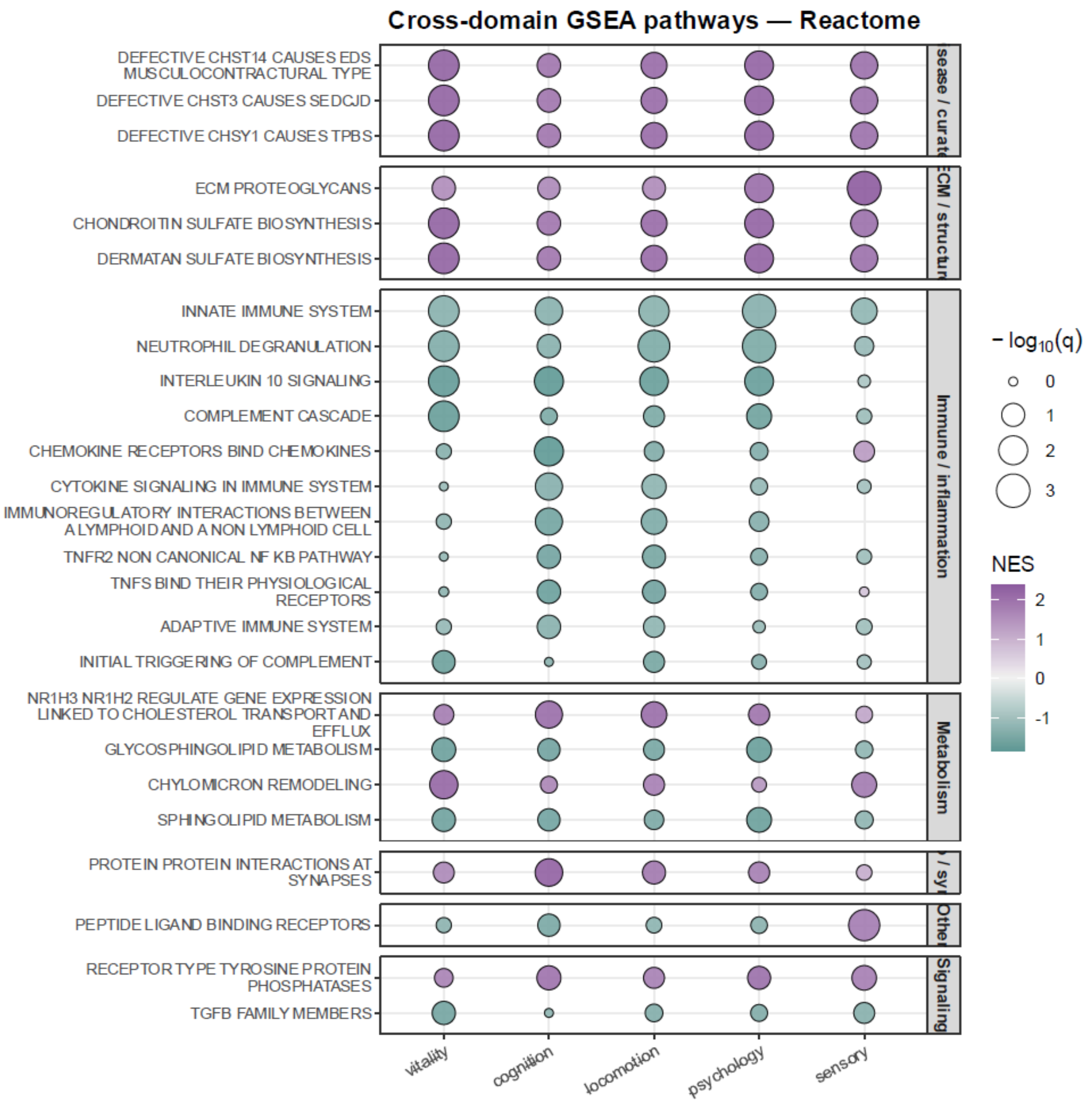


**Supplementary Figure 3. Sex-stratified hazard ratios for the composite IC score across disease, cancer, and mortality endpoints.** Sex-specific hazard ratios (per 1-SD increase in the composite IC score) are shown for (**a**) incident disease endpoints grouped by clinical category, (**b**) cancer incidence endpoints, and (**c**) all cause and cause-specific mortality endpoints. All estimates are from adjusted model (age, sex, ethnicity, Townsend deprivation index, educational qualifications, assessment year and season, smoking status, and alcohol intake frequency). Red circles denote females and blue triangles denote males; horizontal lines indicate 95% confidence intervals; faded points indicate p >= 0.05. Numbers in parentheses show the sex-specific event counts. Sex-specific cancers (breast, ovary, uterine; prostate) appear only in the relevant stratum.

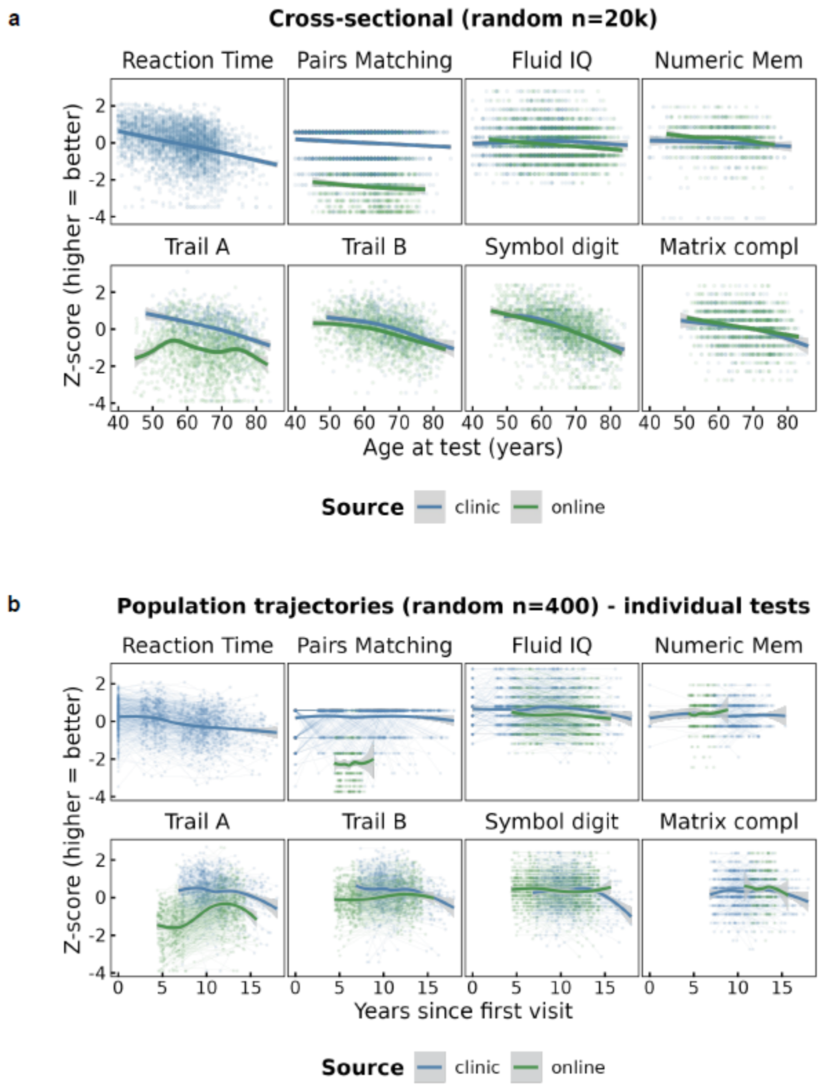


**Supplementary Figure 4. Cross-sectional and longitudinal trajectories of individual cognitive test measures by assessment source.** Individual cognitive test measures contributing to the cognition domain are shown separately to illustrate heterogeneity across the UK Biobank cognitive battery. (**a**) cross-sectional age patterns using a random subset of 20,000 observations, with standardized test scores (Z-scores; higher = better) plotted against age at assessment. (**b**) longitudinal trajectories using a random subset of 400 participants with repeated assessments, with scores plotted against years since first visit; faint lines connect repeated measures within individuals. Smoothed curves summarize the average pattern for clinic-based assessments (blue) and online assessments (green). The battery includes reaction time, pairs matching, fluid intelligence, numeric memory, Trail Making Test A, Trail Making Test B, symbol digit substitution, and matrix pattern completion.
